# Targeting hyperarousal to improve sleep: A network intervention analysis of a digital intervention for insomnia

**DOI:** 10.64898/2026.02.06.26345753

**Authors:** Linda T. Betz, Robert Göder, Gitta A. Jacob, Dieter Riemann

## Abstract

**Objective:** Digital cognitive behavioral therapy for insomnia (CBT-I) is an effective and scalable treatment for chronic insomnia. However, treatment outcomes are typically evaluated using aggregated symptom scores, which obscure differential effects on individual symptoms and limit insights into underlying mechanisms. This study applied network intervention analysis (NIA) to investigate how *somnovia*, a self-guided digital CBT-I intervention, is associated with changes in individual symptoms of insomnia, depression, and anxiety over time.

**Method:** This secondary analysis used data from a randomized controlled trial including 290 adults with chronic insomnia who were randomized to *somnovia* plus treatment as usual or treatment as usual alone. Using item-level data from the Insomnia Severity Index (ISI), Patient Health Questionnaire–9 (PHQ-9), and Generalized Anxiety Disorder–7 (GAD-7) collected at baseline, 3 months, and 6 months, NIA was conducted to identify direct and indirect associations between treatment assignment and individual symptoms.

**Results:** At 3 months, *somnovia* was primarily associated with reductions in sleep-related worry, dissatisfaction with sleep quality, and difficulties relaxing, indicating early effects on cognitive and physiological arousal processes. Improvements in other symptoms appeared to occur indirectly through these core arousal-related processes. By 6 months, treatment-related associations were more broadly distributed across the symptom network, consistent with generalized improvements extending beyond sleep-specific complaints.

**Conclusion:** Together, these findings suggest that *somnovia* exerts its clinical effects by reducing hyperarousal as a central transdiagnostic process, through which improvements in insomnia may contribute to sustained benefits in broader mental health outcomes, including symptoms of depression and anxiety.

**Trial Registration:** ClinicalTrials.gov (NCT05558865).

## Introduction

Approximately 20% of the adult population in Germany report low quality of sleep, and around 6% meet criteria for chronic insomnia (Schlack et al., 2013). Insomnia is linked to substantial impairments in daily functioning and quality of life, particularly if left untreated (Heidbreder et al., 2024). Beyond its standalone clinical relevance, insomnia is a predictor and feature of many other mental disorders, such as depression and anxiety, rendering it an important transdiagnostic treatment target (Baglioni et al., 2011; Hall et al., 2025; Hertenstein et al., 2019; Reesen et al., 2024; Sivertsen et al., 2012).

The first-line treatment for insomnia is cognitive behavioral therapy for insomnia (CBT-I), which integrates behavioral interventions such as sleep restriction with cognitive techniques aimed at modifying maladaptive sleep-related beliefs (Furukawa et al., 2024; Riemann et al., 2017, 2023). When delivered by trained health-care professionals, CBT-I yields large improvements in insomnia severity, with average effect sizes ranging from *d* = 0.84 to 1.27 (Geiger-Brown et al., 2015; Simon et al., 2023; van Straten et al., 2018; Wu et al., 2015). However, the demand for appropriately qualified providers far exceeds their availability (Koffel et al., 2018; Morin, 2015; Riemann et al., 2022). Digital delivery formats of CBT-I therefore represent a scalable alternative and have demonstrated comparable effectiveness, with an average effect size of *d* = 0.78 on insomnia severity (Simon et al., 2023).

Although digital CBT-I interventions have demonstrated robust overall effectiveness, treatment outcomes are typically evaluated using aggregated sum scores. These sum scores treat individual symptoms as interchangeable indicators of an underlying disorder entity (Fried & Nesse, 2015). For example, within the Insomnia Severity Index (ISI), difficulties initiating sleep and impairments in daytime functioning contribute equally to the sum score, despite representing distinct symptom domains that may respond differentially to treatment. While aggregating distinct symptoms into a single total score for analysis is consistent with standard practice in randomized controlled trials (RCTs), aligning with typical regulatory requirements, this approach obscures clinically relevant differences in how individual symptoms respond to treatment (Blanken et al., 2019; Fried & Nesse, 2015). By contrast, shifting the level of analysis to individual symptoms aligns more closely with how clinicians conceptualize and treat psychopathology (Hofmann et al., 2016; Hofmann & Hayes, 2019; McNally, 2021). For example, a patient may fall asleep more quickly following sleep restriction, while improvements in daytime functioning or reductions in sleep-related worry emerge more gradually, reflecting the complementary effects of different CBT-I components (Blanken et al., 2021).

Network Intervention Analysis (NIA) provides a suitable framework for examining intervention effects at the level of individual symptoms (Blanken et al., 2019). NIA is grounded in the *Network Approach to Psychopathology*, which conceptualizes mental disorders as systems of interacting symptoms (Betz et al., 2020; Briganti et al., 2024; Hofmann et al., 2016; McNally, 2021; Robinaugh et al., 2020). Thus, NIA shifts the analytic focus from aggregate outcome measures to symptom-level associations between an intervention and clinical change. By explicitly modeling interrelations among symptoms, this framework allows for the distinction between symptoms that are directly associated with the intervention and those that may change indirectly via connections with other symptoms (Blanken et al., 2019; Fishbein et al., 2023; Lancee et al., 2022). Thus, NIA helps elucidate the processes through which treatment effects of a particular intervention emerge.

The present study focuses on *somnovia*, a fully self-guided, web-based intervention that translates the core components of CBT-I into a structured digital treatment program. Its overall effectiveness has been demonstrated in a recent RCT, showing reductions in insomnia severity, depression, and anxiety, as well as improvements in social and occupational functioning after 3 months of treatment, with effects remaining stable at 6 months (Specht et al., 2024). Building on these findings, the goal of the present secondary analysis is to obtain a more differentiated understanding of how the use of *somnovia* relates to change at the level of individual symptoms. In addition, symptoms of insomnia, depression and anxiety are modeled jointly, reflecting their close clinical interrelations. This modeling approach allows us to explore whether *somnovia* is primarily associated with changes in insomnia symptoms, changes in one or the other domains, or both, which has implications for understanding transdiagnostic effects and for informing preventive strategies through the treatment of insomnia.

## Methods

### Study design and sample

The present secondary analysis is based on data from an RCT evaluating the effectiveness of *somnovia*, a fully self-guided digital CBT-I intervention. The trial was approved by the ethics committee of the Medical Faculty of the University of Kiel, Germany (reference number: D495/22; date: September 5, 2022) and registered at ClinicalTrials.gov (NCT05558865). A total of 290 German-speaking adults with chronic insomnia were recruited online and randomized (1:1) to *somnovia* plus treatment as usual (TAU) or TAU alone. TAU reflected routine care conditions, with no restrictions on concomitant psychotherapy or medication use in both study groups. Sample size in the original trial was determined based on an anticipated effect size of Cohen’s *d* = 0.40 and an expected dropout of 30%.

Inclusion criteria were age ≥ 18 years, a diagnosis of chronic insomnia according to ICD-11 criteria, impaired sleep (ISI total score ≥ 10), and provision of informed consent. The diagnosis of chronic insomnia was established using a structured diagnostic interview conducted by trained psychological staff. No exclusion criteria were applied to enhance ecological validity. The sample had a mean age of 49.8 years (SD = 14.1), and 214 participants (73.8%) were female. Assessments were conducted at baseline, 3 months (T1; primary endpoint), and 6 months (T2; follow-up). The study was conducted in accordance with the Declaration of Helsinki; the privacy rights of human subjects were observed, and informed consent was obtained from all participants. Full details are reported in Specht et al. (2024).

### Measurements

Individual items from the ISI, the Patient Health Questionnaire–9 (PHQ-9) and the Generalized Anxiety Disorder–7 (GAD-7) were used in NIA. For all items, higher scores indicate greater severity.

The ISI assesses insomnia severity as a self-report using seven self-report items on a 5-point scale (0-4) covering sleep initiation, sleep maintenance, early awakenings, dissatisfaction with sleep, daytime interference, quality-of-life impairment, and sleep-related worry (Bastien et al., 2001).

The PHQ-9 is a self-report measure of depressive symptom severity consisting of nine items rated on a 4-point scale (0-3) (Kroenke et al., 2001). The items assess core depressive symptoms, including depressed mood, anhedonia, sleep and appetite disturbances, fatigue, concentration difficulties, psychomotor changes, feelings of worthlessness, and suicidal ideation.

The GAD-7 is a self-report measure of anxiety severity comprising seven items rated on a 4-point scale (0-3) (Spitzer et al., 2006). The items assess symptoms of generalized anxiety, including nervousness, uncontrollable worry, excessive worry, difficulty relaxing, restlessness, irritability, and fear of negative events.

### Intervention

Following clinical guideline recommendations, *somnovia* delivers tailored psychoeducation and psychotherapeutic techniques based on an initial assessment of the user’s sleep patterns, habits, and dysfunctional beliefs.

The behavioral core of the program includes sleep restriction therapy, where bedtimes are adjusted to match actual sleep needs, and sleep hygiene training to eliminate sleep-interfering behaviors. These are supported by lifestyle interventions, including the use of implementation intentions to increase physical activity and “urge surfing” techniques to manage nocturnal alcohol cravings. To address cognitive and physiological arousal, the program provides instructions for relaxation techniques, mindfulness, and gratitude interventions.

Cognitive components target the de-escalation of sleep-related distress by addressing catastrophizing thoughts and rumination. This is achieved through a structured decision tree to evaluate the actionability of worries, alongside a “rumination-stop” protocol that utilizes conditioning and relaxation techniques to interrupt negative thought cycles. For details, see Specht et al. (2024).

The dialogues offered as part of the program can ideally be conducted over a period of five to six weeks. Relaxation techniques and lifestyle changes are then further consolidated afterward. Ideally, the program should be used for a duration of three to six months.

### Statistical Analysis

All statistical analyses were performed using *R*, version 4.5.2. The code for the analyses is available at https://github.com/LindaBetz/somnovia_NIA.

#### Item-level intervention effects

To describe univariate between-group differences in item-level change from baseline to 3 we estimated an ANCOVA model for each symptom item with the T1 item score as outcome, treatment condition as predictor, and the corresponding baseline (T0) item as covariate. For each model, we extracted the group effect estimate and *p*-value. Analogously, item-level effects were assessed at T2.

#### Network intervention analysis (NIA)

NIA was conducted closely following the approach described by Blanken and colleagues (2019). Networks were estimated at the item level, where individual symptoms and the treatment condition (intervention vs. control) served as nodes, and the statistical associations between them were represented as edges. These nodes included items from the ISI, PHQ-9, and GAD-7, though we excluded the sleep-related item of the PHQ-9 (item 3) to avoid topological overlap with the ISI (Blanken et al., 2019). We estimated one network per time point: baseline, after 3 months (T1) and after 6 months (T2).

To ensure that follow-up networks were not influenced by residual baseline differences, we removed baseline variance from each symptom item at T1 and T2 prior to network estimation. Specifically, for each item we regressed the follow-up item score on its corresponding baseline score and used the residuals as baseline-adjusted follow-up values. The baseline-adjusted item values were then used for estimation of the networks at T1 and T2. Baseline networks were estimated using the unadjusted baseline item scores. This baseline-adjustment strategy is consistent with the primary data-analytic approach prespecified for the RCT, in which treatment effects on questionnaire sum scores were estimated using baseline-adjusted models (Specht et al., 2024); in the present analyses, this strategy was extended to the item level.

Networks were estimated as mixed graphical models, a special type of partial correlation network which allows for the inclusion of both categorical and continuous variables (Haslbeck & Waldorp, 2020). To obtain a sparse and interpretable edge set, we applied regularization using an L1-penalty (LASSO), which shrinks small associations toward zero (Epskamp & Fried, 2018). The strength of regularization is controlled by a tuning parameter, which was selected via 10-fold cross-validation, following prior work using NIA (Blanken et al., 2019; Lancee et al., 2022). This procedure balances model complexity against goodness of fit and reduces overfitting. To further obtain a conservative edge set, we used the AND-rule, retaining an edge only if it was selected in both corresponding neighborhood regressions. Given that the estimation requires complete cases, the analytic sample size varied slightly across time points (Baseline: N = 290, T1: N = 248, T2: N = 244).

Networks were visualized using the *R* package *qgraph* (Epskamp et al., 2012). In these networks, edges between symptom nodes indicate statistical associations between individual symptoms after controlling for all other symptoms in the network (i.e., a conditional dependence relation). Connections between the treatment node and symptom nodes indicate associations between treatment assignment and specific symptoms, reflecting which symptoms are most directly related to the intervention within the overall modeled symptom system. Since treatment assignment was randomized, associations involving the treatment node can be interpreted as directionally originating from the intervention toward symptoms (i.e., item-specific intervention connections). Importantly, the absence of a direct connection between the treatment node and a given symptom does not imply that the symptom is unaffected by the intervention; rather, it suggests that any association with treatment may be indirect and operate through other symptoms in the network.

To facilitate visual comparison across time points, we computed an average layout of the three estimated networks and plotted each network using this common layout. The positioning of the nodes was determined using the Fruchterman-Reingold algorithm, placing more strongly connected nodes at the center and less connected nodes to the periphery. For readability, very small edges were suppressed using a minimum edge weight threshold of 0.01, and edge weight scaling was set to be comparable across follow-up networks.

We evaluated the accuracy of estimated edge weights by creating 500 bootstrap samples for each estimated network and fitting the model in each of the bootstrapped samples. This procedure provides the sampling distribution for each edge weight. For further details, see Epskamp, Borsboom, & Fried (2017).

## Results

### Item-level intervention effects

After 3 months, *somnovia* showed significant effects on all assessed insomnia, depressive, and anxiety symptoms (see Table 1). After 6 months, effects remained significant for all core insomnia symptoms except *noticeability of impaired quality of life to others* and *difficulty initiating sleep*; for these symptoms, effect estimates were in favor of the intervention group but did not reach statistical significance. Effects on depression and anxiety were largely maintained; symptoms that did not reach statistical significance (suicidal ideation, restlessness, and fear) showed effect estimates in the expected direction, with most *p* values close to the conventional significance threshold. For a visualization of changes in individual symptom scores across time, see Supplementary Figure 1.

**Table 1.**
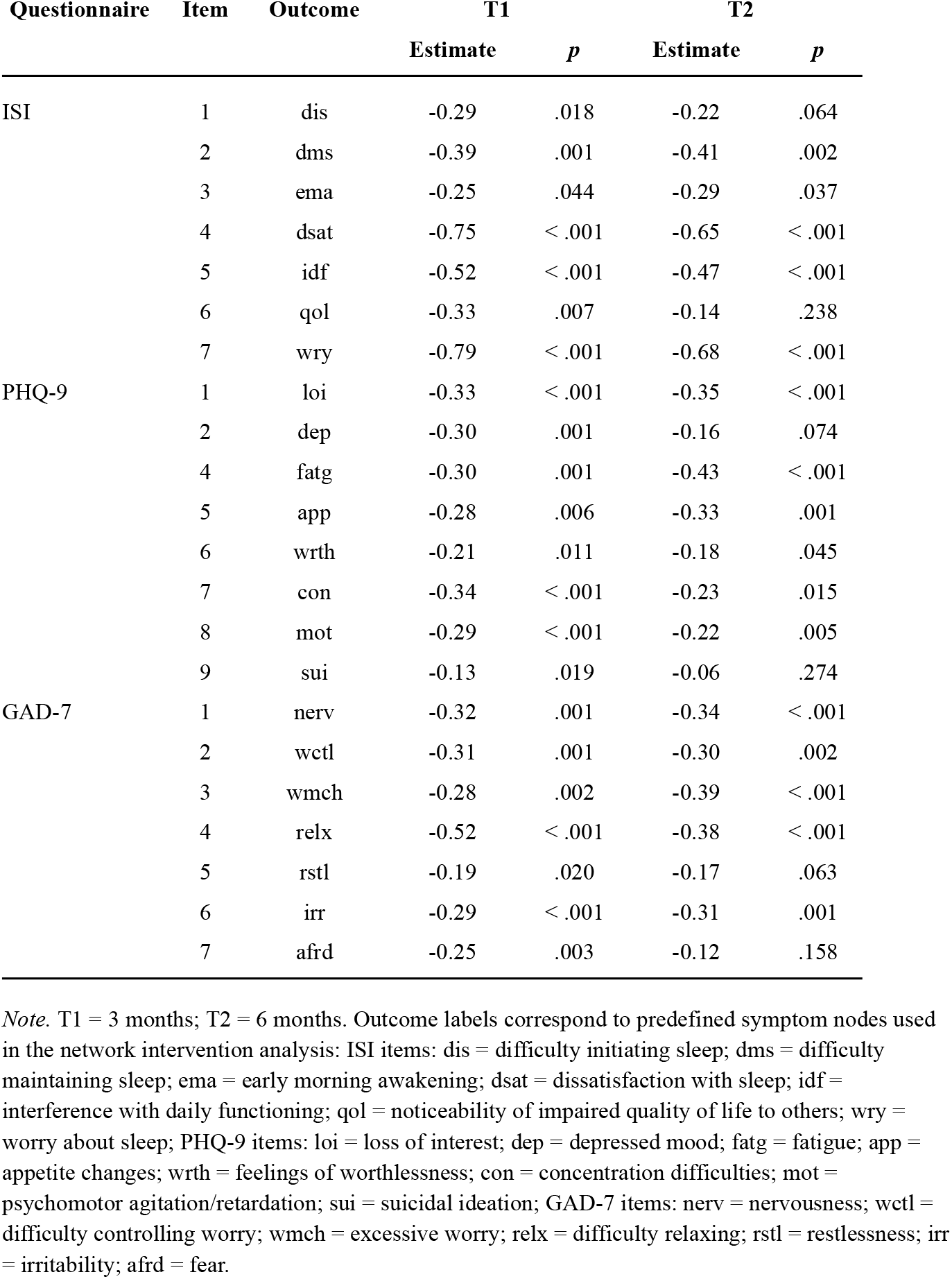
Item-level intervention effects at 3 months (T1) and 6 months (T2) Estimates represent group effects (intervention vs. control). Negative values indicate greater symptom reduction in the intervention group.

### Network Intervention Analysis

As expected, given randomization prior to treatment initiation, NIA showed that at baseline there were no associations between the treatment variable and any symptom in the network (see Figure 1a).

**Figure 1.**
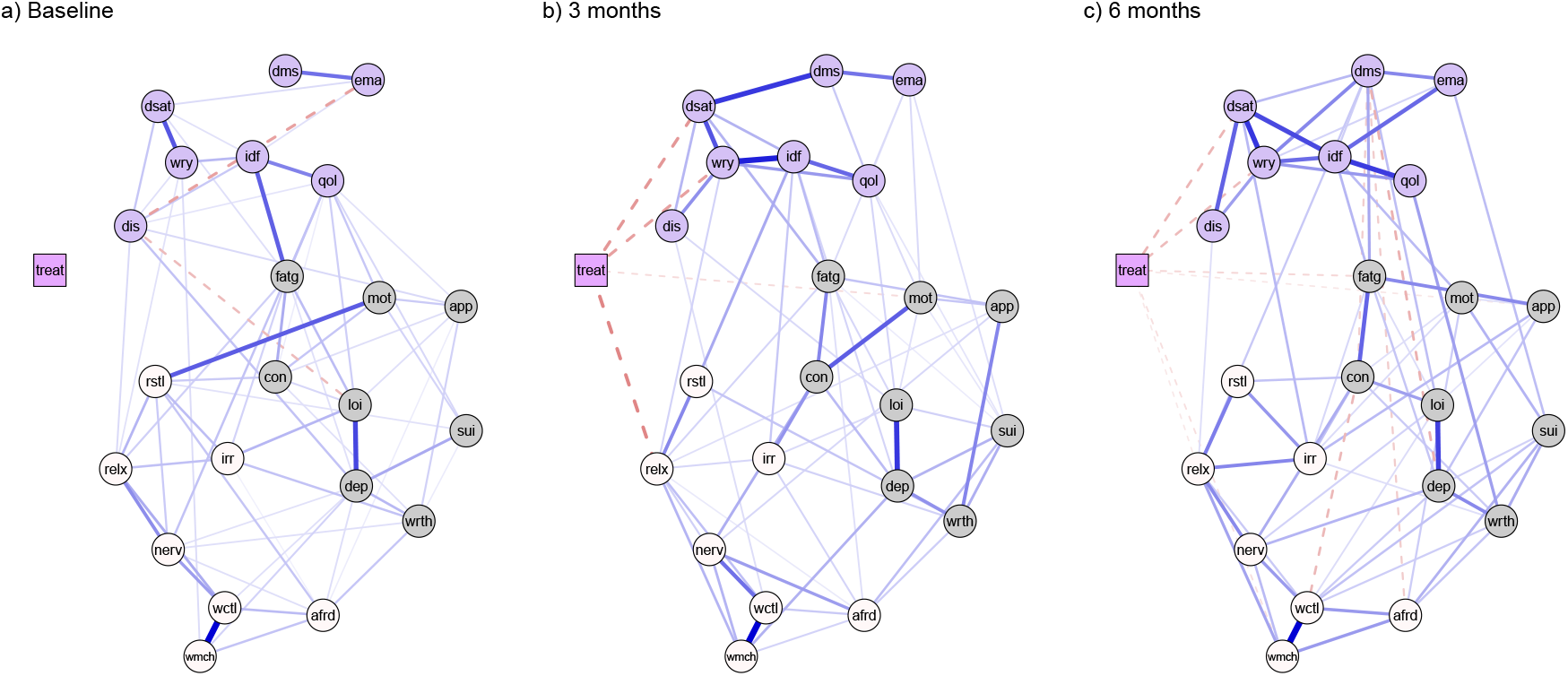
Estimated symptom networks based on Network Intervention Analysis (NIA) at three time points: (a) baseline (pre-treatment), (b) 3 months, and (c) 6 months. Networks include the treatment variable (square), representing assignment to somnovia plus treatment as usual (TAU) versus TAU alone, and symptom variables (circles). Insomnia symptoms assessed with the Insomnia Severity Index (ISI; purple), depressive symptoms assessed with the Patient Health Questionnaire–9 (PHQ-9; grey; sleep item excluded), and anxiety symptoms assessed with the Generalized Anxiety Disorder–7 (GAD-7; beige) are shown. Solid blue lines indicate positive associations, whereas dashed red lines indicate negative associations; dashed red lines connecting the treatment node to symptoms indicate symptom reductions associated with the intervention. *Abbrevations:*treat = treatment (*somnovia* vs. control); ISI items: dis = difficulty initiating sleep; dms = difficulty maintaining sleep; ema = early morning awakening; dsat = dissatisfaction with sleep; idf = interference with daily functioning; qol = noticeability of impaired quality of life to others; wry = worry about sleep; PHQ-9 items: loi = loss of interest; dep = depressed mood; fatg = fatigue; app = appetite changes; wrth = feelings of worthlessness; con = concentration difficulties; mot = psychomotor agitation/retardation; sui = suicidal ideation; GAD-7 items: nerv = nervousness; wctl = difficulty controlling worry; wmch = excessive worry; relx = difficulty relaxing; rstl = restlessness; irr = irritability; afrd = fear.

After 3 months (T1), NIA showed symptom-specific associations of somnovia with the GAD-7 item *difficulty relaxing* (relx; 0.18), the ISI items *dissatisfaction with sleep quality* (dsat; 0.16) and *worry about sleep* (wry; 0.15), as well as a smaller association with the PHQ-9 item *psychomotor agitation/retardation* (mot; 0.06; see Figure 1b). No other symptom-specific associations with treatment assignment were observed at this time point, suggesting that treatment effects on remaining symptoms may operate indirectly through changes in these core symptoms.

At 6 months (T2), NIA showed several associations between treatment assignment and individual symptoms, which varied in strength (see Figure 1c). The strongest associations were observed with the ISI items *dissatisfaction with sleep quality* (dsat; 0.11) and *worry about sleep* (wry; 0.10), indicating that these symptoms remained most closely related to treatment assignment at follow-up. In addition, small associations were present between treatment assignment and other symptoms from the PHQ-9 and GAD-7, namely *fatigue* (fatg; 0.06), *excessive worry* (wmch; 0.05), *difficulty relaxing* (relx; 0.04), and *appetite changes* (app; 0.04).

Notably, bootstrapping procedures indicated that these treatment-symptom associations were overall robustly identified, as evidenced by high edge inclusion frequencies and non-zero estimates across samples (see Supplementary Figure 2).

## Discussion

In this study, we applied NIA to examine symptom-level effects of *somnovia*, a self-guided, web-based intervention for adults with chronic insomnia (Specht et al., 2024). By moving beyond aggregated sum scores and focusing on individual symptoms and their interrelations, this approach allows for a more nuanced investigation of how treatment effects develop within and across symptom domains over time (Blanken et al., 2019; Briganti et al., 2024; Fried & Nesse, 2015).

At 3 months, *somnovia* showed direct associations with a small set of symptoms, including difficulty relaxing, dissatisfaction with sleep quality, worry about sleep, and, to a smaller extent, psychomotor complaints. This pattern suggests that at this treatment phase, the effects of *somnovia* are concentrated on hyperarousal-related processes that operate across both nighttime and daytime functioning (Dressle & Riemann, 2023; Harvey, 2002; Palagini et al., 2015; Riemann et al., 2010): Whereas the associations with reduced sleep-related worry and dissatisfaction with sleep quality may represent changes in cognitive-emotional arousal, the links with difficulty relaxing and psychomotor complaints may point to reductions in more physiologically anchored aspects of hyperarousal. By reducing these interrelated aspects of hyperarousal in parallel, *somnovia* may foster a calmer, less vigilant engagement with sleep, thereby weakening the vicious cycle of arousal and frustration that perpetuates insomnia (Dressle & Riemann, 2023; Espie et al., 2014).

Importantly, this focused pattern of direct treatment-symptom associations should not be interpreted as indicating that *somnovia* affects only a limited subset of symptoms. As shown in the univariate item-level analyses, improvements were observed across all symptoms. Conversely, NIA suggests that these broader effects are likely driven indirectly by changes in arousal-related core symptoms. The observed symptom-symptom associations support this interpretation: Reduced sleep-related worry, for example, was linked to fewer difficulties initiating sleep and less interference with daily functioning, consistent with the observation that lower cognitive arousal facilitates sleep onset (Wuyts et al., 2012).

Our results are broadly consistent with previous findings based on NIA. Compared with Blanken et al. (2019), who examined symptom-level effects of a digitally delivered CBT-I intervention with human support over a 10-week period, we also observed direct effects on cognitive symptoms such as worry about sleep and dissatisfaction with sleep quality. In contrast, the pronounced effects on difficulties maintaining sleep reported at week 10 in their study were not evident in our 3-month network. In addition to temporal variations in when symptom-level effects become observable, these differences are not necessarily indicative of inconsistent findings. Rather, they may reflect the fact that, despite sharing a common CBT-I framework, individual interventions may emphasize different therapeutic components and, therefore, mechanistic targets (Furukawa et al., 2024). Beyond the core content, the specific intervention design, including the structural delivery (e.g., interactive dialogues vs. module-based navigation), narrative tone, and the integration of modalities such as visual illustrations or audio-guided exercises, may further influence how users process and internalize therapeutic techniques. Such differences are in turn expected to manifest at the symptom level.

Further conceptual support for this interpretation comes from Blanken et al. (2021), who identified distinct symptom-specific effects of behavioral therapy (BT) and cognitive therapy (CT) components within CBT-I. Specifically, BT was uniquely associated with improvements in sleep efficiency and dissatisfaction with sleep, whereas CT showed stronger links to cognitive and daytime-related symptoms, including worry about sleep and interference with daily functioning. As CBT-I combines cognitive and behavioral components, the presence of symptom-level effects spanning both domains is theoretically expected in *somnovia*. Moreover, behavioral components of CBT-I have been shown to produce relatively rapid effects, whereas effects of CT tend to unfold slower and last longer (Harvey et al., 2014). From this perspective, the prominence of cognitive and arousal-related symptoms in our 3-month network is theoretically plausible.

At 6 months, the pattern of associations changed. While dissatisfaction with sleep quality and worry about sleep remained the symptoms most strongly associated with treatment assignment, additional, weaker associations emerged with other symptoms, including difficulties relaxing, excessive worrying, fatigue, appetite, and psychomotor complaints. Importantly, these additional associations were small and partially unstable, warranting cautious interpretation. Nevertheless, the overall pattern suggests a transition from a focused, symptom-specific treatment impact toward a more distributed influence across the symptom network. This finding is generally in line with previous NIA findings, where effects of CBT-I on individual symptoms became more dispersed over time (Blanken et al., 2019). Earlier in treatment, interventions may primarily affect a limited number of symptoms directly targeted by the therapeutic components. As these core symptoms improve, their influence may “spread” through the network, leading to secondary improvements in interconnected symptoms over time (Borsboom, 2017). From this perspective, the dispersion of direct treatment-symptom associations at follow-up should not be interpreted as diminishing treatment effectiveness. Rather, it may reflect a redistribution of treatment-related variance across the network, with benefits increasingly embedded in the broader symptom system rather than tied to isolated symptoms. In this sense, the intervention may increasingly function as a broad stabilizing factor within the symptom network, supporting more generalized symptom improvement rather than having isolated effects on specific complaints (Borsboom, 2017; Robinaugh et al., 2020).

From a transdiagnostic perspective, the present findings suggest that insomnia, and hyperarousal specifically, functions as an upstream intervention target whose effects on broader mental health outcomes unfold indirectly over time. At 3 months, no direct treatment-symptom associations were observed for core symptoms of depression or anxiety, suggesting that early effects of *somnovia* on these outcomes are mediated through changes in insomnia-centered hyperarousal rather than reflecting direct intervention effects. By 6 months, a broader pattern of symptom associations emerged, consistent with more generalized improvements extending beyond sleep-specific complaints. Taken together, these findings support the view that sleep-related symptoms, and hyperarousal specifically, represent important transdiagnostic intervention targets within a broader mental health symptom network (Hall et al., 2025; Harvey, 2009; Hertenstein et al., 2019, 2022; Riemann et al., 2020). Targeting insomnia in this manner may therefore initiate beneficial effects that extend beyond sleep itself and contribute to longer-term improvements in depression and anxiety (Reesen et al., 2024).

### Strengths and Limitations

A major strength of this study is its comparatively large sample size, which enabled stable estimation of NIA. Moreover, the broad assessment of psychopathology allowed us to move beyond sleep-specific outcomes and examine how an insomnia intervention influences a wider, transdiagnostic symptom network. Several limitations should nevertheless be acknowledged. First, this analysis was based on pre-existing trial data, which constrained the temporal resolution of symptom assessments to two follow-up time points after 3 and 6 months. While informative, this design does not capture the fine-grained temporal dynamics that more frequent (e.g., weekly) measurements could provide, and thus necessarily limits our ability to characterize the unfolding of intervention effects. Second, outcomes were based exclusively on self-report measures; although appropriate for subjective sleep-related experiences, future studies could benefit from incorporating objective indicators such as actigraphy. Finally, because *somnovia* is a multi-component intervention, the relative contributions to symptom-level effects of its behavioral and cognitive elements cannot be disentangled. Comparative designs are needed to elucidate the mechanisms driving changes in individual symptoms through different interventional approaches (Blanken et al., 2021; Lancee et al., 2022).

### Conclusion

In conclusion, this study underscores that NIA can provide valuable insights into symptom-specific treatment pathways, allowing for a more nuanced understanding of the mechanisms through which the intervention unfolds. Applying this approach revealed that *somnovia* exerts its effects primarily by reducing cognitive and physiological arousal. From a transdiagnostic perspective, targeting these core processes may initiate broader improvements across interconnected symptom domains, thereby supporting reductions in emotional distress and functional impairment beyond sleep-specific outcomes.

## Data Availability

Due to proprietary restrictions, the dataset analyzed in this manuscript are not publicly available but are available from the corresponding author upon request.

## Required statements

## Acknowledgements

The authors would like to thank all individuals who participated in the *somnovia* trial.

## CRediT authorship contribution statement

LTB: Conceptualization, Formal analysis, Visualization, Writing – original draft, Writing – review & editing; RG: Investigation, Writing – review & editing; GAJ: Investigation, Project administration, Writing – original draft, Writing – review & editing; DR: Writing – review & editing.

## Declaration of competing interest

*GAIA*, the developer of *somnovia*, funds the positions of LTB and GAJ.

DR is now speaker of the board of the German Sleep Society (non profit organization) and receives monthly honoraria for this task. DR is a member of the Executive Board of FAVT (Freiburger Ausbildungsinstitut für Verhaltentherapie/Freiburg Institute for Behavioral Therapy; non profit organisation). In this function he receives honoraria for running examinations, giving lectures, attending board meetings and participating in the selection of candidates. DR was Editor-in-Chief of the Journal of Sleep Research which is owned by the European Sleep Research Society (non profit organisation) – he received monthly payments for this task till December 2025. DR receives royalties for authored books and book chapters from several publishing companies (Elsevier, Hogrefe, Kohlhammer. Wiley and Sons, etc.). The published materials mainly deal with insomnia and its treatment. DR frequently lectures at conferences, meetings, seminars, mostly invited by the organizing bodies – sometimes honoraria are paid for his engagement and usually travel costs (if travelling is involved) are covered. Most of his talks deal with aspects of insomnia. In the last 36 months DR received lecturing honoraria also from Novartis, GAIA AG and Idorsia. DR receives honoraria from GAIA (Germany), Meinstresscoach (Switzerland), /Mind (Germany), 7Mind (Germany), HelloBetter (Germany), Sympatient (Germany) and X-Trodes (Israel) for advising on the development of internet-based approaches to insomnia treatment and development of new measurement devices.

RG has no competing interests to declare.

## Declaration of generative AI use

During the preparation of this work, the authors used Gemini in order to assist with formatting and to support checks for compliance with journal guidelines. After using this tool, the authors reviewed and edited the content as needed and take full responsibility for the content of the published article.

## Funding statement

This work was supported by GAIA, the developer, owner, and manufacturer of *somnovia*.

## Notes

### Clinical Trial

NCT05558865

### Author Declarations

The ethics committee of the Medical Faculty of the University of Kiel, Germany gave ethical approval for this work (reference number: D495/22; date: September 5, 2022).

