## Supplementary Material for "Targeting hyperarousal to improve sleep: A network intervention analysis of a digital intervention for insomnia"

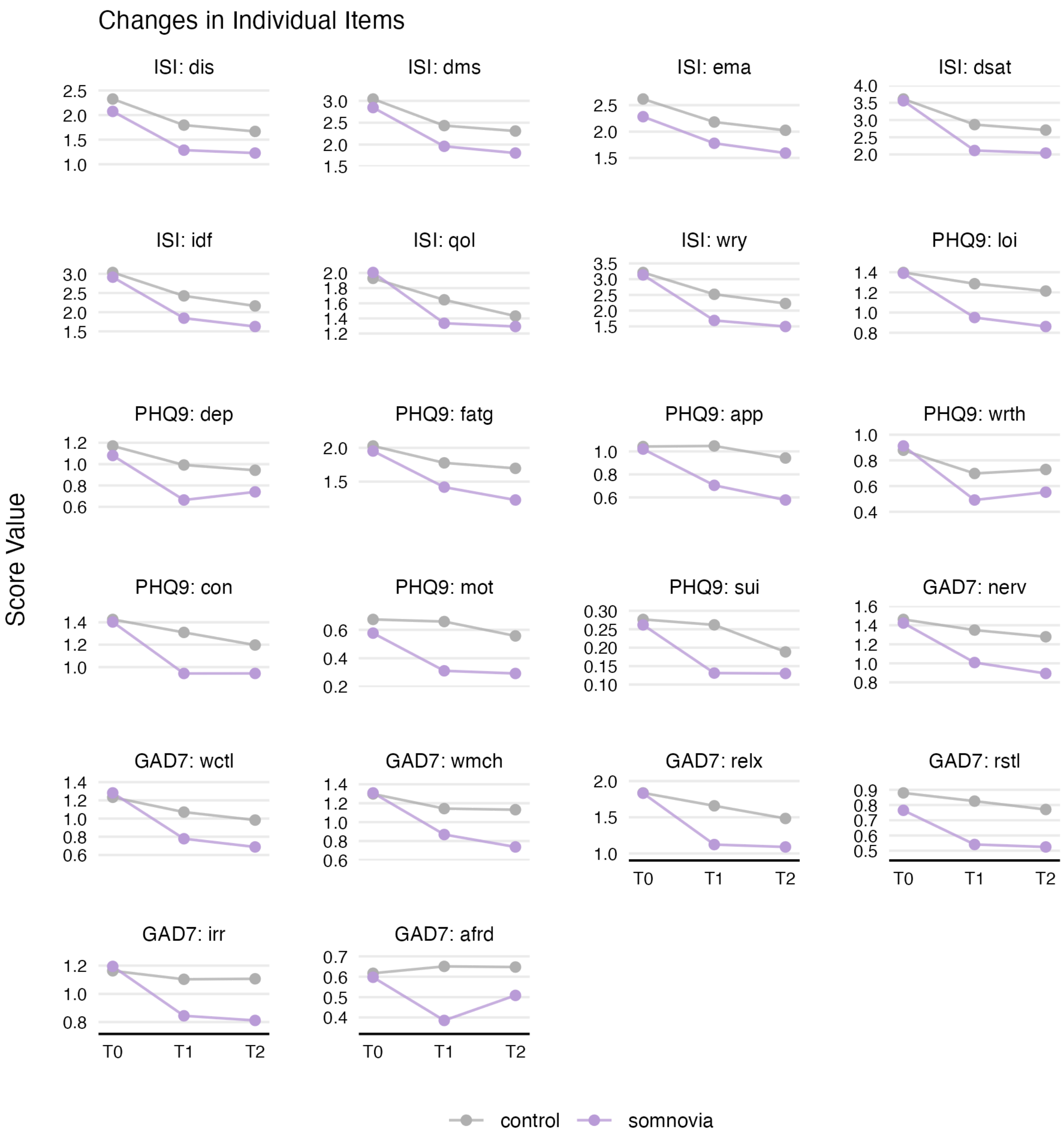


**Supplementary Figure 1.** Longitudinal changes in individual symptom scores for *somnovia* vs. control groups. Mean score values are displayed across three time points: T0 (baseline, pre-treatment), T1 (3 months), and T2 (6 months) for items across the Insomnia Severity Index (ISI), Patient Health Questionnaire–9 (PHQ-9), and Generalized Anxiety Disorder–7 (GAD-7).

*Abbrevations:* ISI items: dis = difficulty initiating sleep; dms = difficulty maintaining sleep; ema = early morning awakening; dsat = dissatisfaction with sleep; idf = interference with daily functioning; qol = noticeability of impaired quality of life to others; wry = worry about sleep; PHQ-9 items: loi = loss of interest; dep = depressed mood; fatg = fatigue; app = appetite changes; wrth = feelings of worthlessness; con = concentration difficulties; mot = psychomotor agitation/retardation; sui = suicidal ideation; GAD-7 items: nerv = nervousness; wctl = difficulty controlling worry; wmch = excessive worry; relx = difficulty relaxing; rstl = restlessness; irr = irritability; afrd = fear.

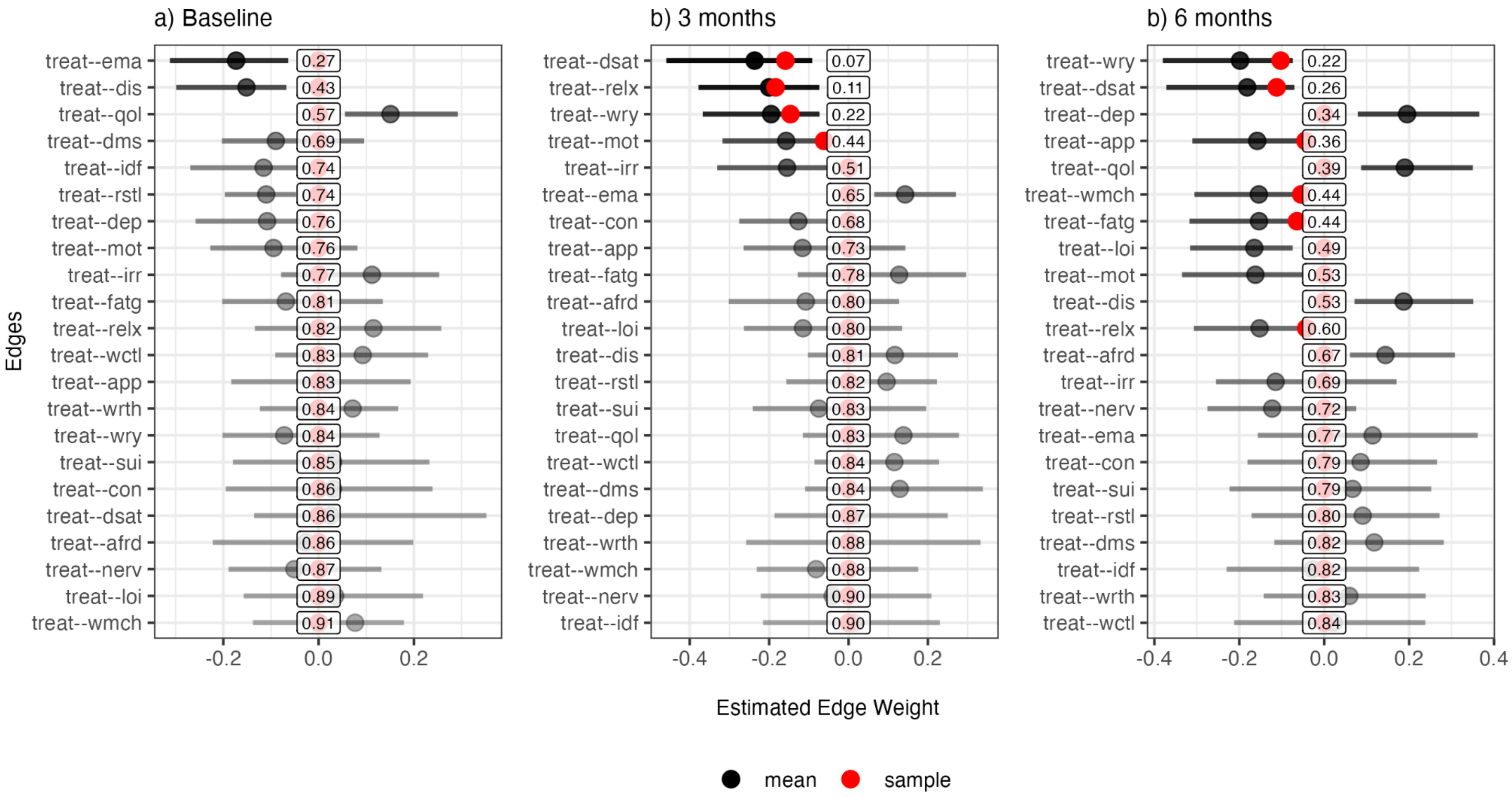
a) Baseline
treat--ema 0.27
treat--dis 0.43
treat--qol 0.57
treat--dms 0.69
treat--idf 0.74
treat--rstl 0.74
treat--dep 0.76
treat--mot 0.76
treat--irr 0.77
treat--fatg 0.81
treat--relx 0.82
treat--wctl 0.83
treat--app 0.83
treat--wrth 0.84
treat--wry 0.84
treat--sui 0.85
treat--con 0.86
treat--dsat 0.86
treat--afrd 0.86
treat--nerv 0.87
treat--loi 0.89
treat--wmch 0.91
-0.2
0.0
0.2
b) 3 months
treat--dsat 0.07
treat--relx 0.11
treat--wry 0.22
treat--mot 0.44
treat--irr 0.51
treat--ema 0.65
treat--con 0.68
treat--app 0.73
treat--fatg 0.78
treat--afrd 0.80
treat--loi 0.80
treat--dis 0.81
treat--rstl 0.82
treat--sui 0.83
treat--qol 0.83
treat--wctl 0.84
treat--dms 0.84
treat--dep 0.87
treat--wrth 0.88
treat--wmch 0.88
treat--nerv 0.90
treat--idf 0.90
-0.4
-0.2
0.0
0.2
b) 6 months
treat--wry 0.22
treat--dsat 0.26
treat--dep 0.34
treat--app 0.36
treat--qol 0.39
treat--wmch 0.44
treat--fatg 0.44
treat--loi 0.49
treat--mot 0.53
treat--dis 0.53
treat--relx 0.60
treat--afrd 0.67
treat--irr 0.69
treat--nerv 0.72
treat--ema 0.77
treat--con 0.79
treat--sui 0.79
treat--rstl 0.80
treat--dms 0.82
treat--idf 0.82
treat--wrth 0.83
treat--wctl 0.84
-0.4
-0.2
0.0
0.2
0.4
Edges
Estimated Edge Weight
mean
sample

**Supplementary Figure 2.** Network stability and edge weight estimates for treatment assignment across study phases. The panels display edge weights representing the relationship between treatment assignment (*somnovia* vs. control) and symptom nodes at (a) Baseline (pre-treatment), (b) 3 months, and (c) 6 months, derived from network intervention analysis (NIA). Shaded areas represent 95% confidence intervals (CIs) obtained via bootstrapping; these CIs are conditioned on edge presence and are calculated only for networks in which the edge was included (non-zero). CI transparency denotes the selection frequency (stability) of the edge across bootstrap samples, while the numerical value within each box indicates the proportion of samples in which the edge was excluded (sparsity).

*Abbreviations:* treat, treatment (*somnovia* vs. control); ISI items: dis = difficulty initiating sleep; dms = difficulty maintaining sleep; ema = early morning awakening; dsat = dissatisfaction with sleep; idf = interference with daily functioning; qol = noticeability of impaired quality of life to others; wry = worry about sleep; PHQ-9 items: loi = loss of interest; dep = depressed mood; fatg = fatigue; app = appetite changes; wrth = feelings of worthlessness; con = concentration difficulties; mot = psychomotor agitation/retardation; sui = suicidal ideation; GAD-7 items: nerv = nervousness; wctl = difficulty controlling worry; wmch = excessive worry; relx = difficulty relaxing; rstl = restlessness; irr = irritability; afrd = fear.
